# Neura: a specialized large language model solution in neurology

**DOI:** 10.1101/2024.02.11.24302658

**Authors:** Sami Barrit, Nathan Torcida, Aurélien Mazeraud, Sébastien Boulogne, Jeanne Benoit, Timothée Carette, Thibault Carron, Bertil Delsaut, Eva Diab, Hugo Kermorvant, Adil Maarouf, Sofia Maldonado Slootjes, Sylvain Redon, Alexis Robin, Sofiène Hadidane, Vincent Harlay, Vito Tota, Tanguy Madec, Alexandre Niset, Salim El Hadwe, Nicolas Massager, Stanislas Lagarde, Romain Carron

## Abstract

Large language models’ (LLM) ability in natural language processing holds promise for diverse applications, yet their deployment in fields such as neurology faces domain-specific challenges. Hence, we introduce Neura: a scalable, explainable solution to specialize LLM. Blindly evaluated on a select set of five complex clinical cases compared to a cohort of 13 neurologists, Neura achieved normalized scores of 86.17% overall, 85% for differential diagnoses, and 88.24% for final diagnoses (55.11%, 46.15%, and 70.93% for neurologists) with rapid response times of 28.8 and 19 seconds (9 minutes and 37.2 seconds and 8 minutes and 51 seconds for neurologists) while consistently providing relevant, accurately cited information. These findings support the emerging role of LLM-driven applications to articulate human-acquired and integrated data with a vast corpus of knowledge, augmenting human experiential reasoning for clinical and research purposes.

## 1 Introduction

Artificial Intelligence (AI) has become an instrumental force across multiple sectors, notably in healthcare (Yu et al., 2018) and research (Dong et al., 2021). Within this expansive realm, large language models (LLM) have garnered attention for their proficiencies in natural language processing (NLP). These models have demonstrated versatility in diverse broad applications, most recently exemplified by the advent of conversational agents (Radford et al., 2018; Devlin et al., 2018; Brown et al., 2020). However, their deployment in specialized scientific domains, particularly medicine, is distinctly challenging (Beam et al., 2023), due to the stringent constraints inherent to medical applications and the nuanced, discipline-specific considerations such domains entail (Ling et al., 2023). Neurology —with its intricate clinical manifestations, neural substrates, and interdisciplinary integration— is a prime example of a complex and rapidly evolving expanse of knowledge that may be substantially embedded —and effectively encoded— in natural language.

Hence, a salient challenge resides in fine-tuning LLM to achieve domain-specific relevance. Traditional fine-tuning methods are resource-intensive, requiring substantial computational and human capital (Strubell et al., 2019). Consequently, these methods are often feasible only for large-scale projects with considerable resources (Singhal et al., 2022). Another limitation of conventional LLM implementation is interpretability and transparency in information processing (Holzinger et al., 2017; Lipton, 2018) — a critical requirement for verifiable information generation for medical and research purposes. Furthermore, prevalent LLM are often constrained by token-based context limitations, which restrict their utility in complex, data-rich environments typical of healthcare and research (Huang et al., 2019). Here, we introduce a scalable, verifiable, and hypercontextual solution to specialize LLM, aligning with the principles of explainable artificial intelligence (XAI) (Doshi-Velez and Kim, 2017) (Figure 1). Then, we study its diagnostic performance compared to neurologists in complex case scenarios mimicking clinical practice.

**Figure 1.**
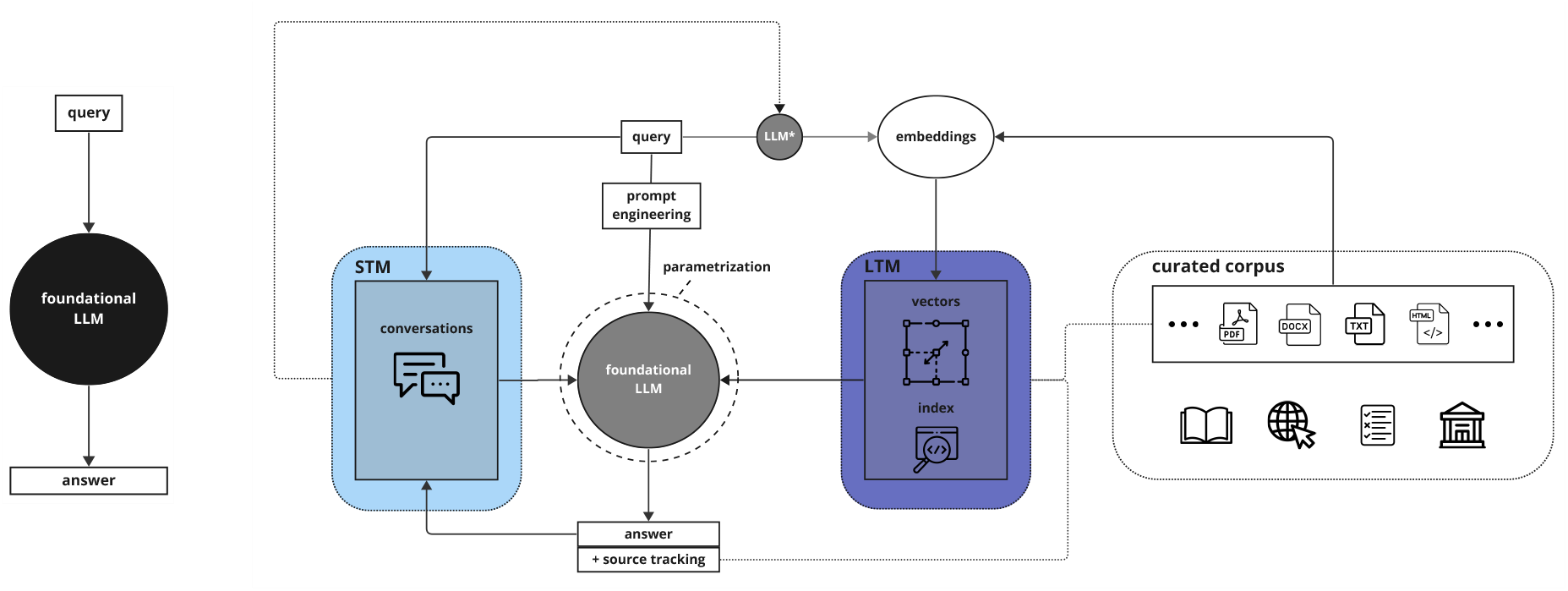
(left) black-box use of LLM; (right) Neura solution’s architecture. LLM: large language models, *fine-tuned LLM for embeddings generation; LTM: long-term memory; STM: short-term memory

## 2 Methods

### 2.1 Neura — a specialized LLM

Neura is a solution deploying LLM with custom parameters and prompt engineering on curated corpora with extended contexts (Sciense, New York, US) for advanced retrieval augmented generation (Lewis et al., 2020). This solution is predicated on a dual-database architecture integrating both ‘long-term memory’ (LTM) and ‘short-term memory’ (STM) components. The LTM serves as the repository for domain-specific knowledge. It employs an agnostic, vectorized approach enabled by text embeddings generated from parsed source texts (Mikolov et al., 2013). The STM captures the setting and conversational history between the user and the LLM, thereby adding a layer of contextual knowledge. The STM is implemented using a non-relational database (Pokorny, 2011), ensuring real-time accessibility and state persistence of conversational data. Information retrieval is optimized in speed and accuracy through a single-stage filtering process, integrating vector and metadata indexes into a unified structure (Taipalus, 2023). Source tracking is enabled, culminating in actionable, standardized references for the end-user and ensuring verifiability of answer accuracy. For this study, we deployed a state-of-the-art LLM, GPT-4 Turbo (OpenAI, San Francisco, US), on a prototype corpus curated for clinical neurology sourced from five comprehensive neurology textbooks (Samuels et al., 2014; Jankovic et al., 2021; Campbell and DeJong, 2005; Merritt, 2010), the neurologic disorders section of Merck’s Manual (copyrighted) (MSD, 2024), and Wikipedia (open-source) (Wikipedia, 2024) (Figure 1).

### 2.2 Diagnostic challenges

Five representative cases were adapted from Neurology’s Resident & Fellow Clinical Reasoning section (Francis et al., 2015; Choi et al., 2017; Harada et al., 2019; Lun et al., 2020; McIntosh and Scott, 2021) to mirror the clinical practice through a two-tiered diagnostic approach. The first tier required formulating and justifying an exhaustive differential diagnosis based on initial clinical presentation and findings. In the second tier, conclusive clinical information was provided to establish a definitive diagnosis. We recruited senior residents and board-certified neurologists from teaching hospitals in Belgium and France. Neurologists engaged in complex clinical reasoning to solve these diagnostic challenges, solely relying on intrinsic knowledge in the first tier — external resources were permitted in the second tier. All challenges were conducted via videoconferencing sessions, supervised by two investigators who provided documents presenting the cases, initial instructions, and procedural assistance. Answers with timing were recorded in text documents, which were subsequently collected and anonymized. Neura undertook the challenges based on the same documents provided to neurologists. Two senior academic neurologists, each responsible for residency training and educational programs at their respective universities, independently evaluated the answers, blinded to the involvement of Neura as a participant. They employed a standardized scoring sheet derived from the published corrections of the cases, assigning points for precise and justified diagnoses and allowing bonus points for unexpected, relevant findings. Incorrect or risky conclusions incurred deductions, with a two-point loss yielding a null question score. Null scores from both evaluators constituted question failure. If multiple participants achieved the maximum score for a given question, the evaluator chose a preferred answer; conversely, if a single answer attained the maximum score, it was then defined as the highest score. In parallel, an independent investigator assessed the reliability and verifiability of the AI-generated information. This was achieved by first classifying the references provided within the answers as relevant, irrelevant, or hallucinatory (i.e., incorrect or nonexistent), then ensuring all generated information was accurately derived from the cited sources, resulting in a binary outcome (accurate or inaccurate).

### 2.3 Statistical analysis

Descriptive statistics were calculated for the scores and times. For normalization, the maximum possible combined score for each question was determined by summing the highest score assigned by each evaluator. For any participant, we calculated the combined score from both evaluators for each question and then divided this by its maximum possible combined score. These resulting normalized scores were expressed as percentages, indicating the proportion of the maximum possible points each participant collected on a given question. We used the intraclass correlation coefficient (ICC) to measure consistency agreement for inter-rater reliability between evaluators. We compared the performance of Neura with that of neurologists using a linear mixed-effects model. Before analysis, we used residual plots, QQ plots, and Shapiro-Wilk tests to assess the assumptions of normality, homoscedasticity, and random effects structure. This model utilized average scores derived from the two evaluators as the dependent variable. Participant type (Neura vs. human) was treated as a fixed effect, while variability across questions was modeled as random. The significance of the fixed effect was corroborated using an ANOVA with Satterthwaite’s method for approximating degrees of freedom. We employed a Monte Carlo simulation (MCS) of 10,000 iterations to estimate the probabilities for Neura achieving observed thresholds of maximum scores, highest scores, and preferred answers among its 20 scores by chance — assuming a uniform distribution of scores within each question’s specific range across all participants. We set our alpha level threshold at 0.05 to determine statistical significance using two-tailed tests. All computations and visualizations were performed using R version 4.1.3, with the packages: ‘afex,’ ‘eulerr,’ ‘ggplot2,’ ‘irr,’ ‘lme4’, and ‘lmertest.’ Anonymized data not published within this article will be made available by request from any qualified investigator.

## 3 Results

Of the 13 neurologists, 8 were board-certified. Challenges were conducted between March and October 2023. ICC(C,2) was found to be significant at 0.767 (95% CI [0.675, 0.833], F(139,139) = 4.3, p < 0.001). The residuals did not significantly deviate from normality (W = 0.99327, p = 0.753, Shapiro-Wilk test) as observed on the QQ plot, and plots of residuals versus fitted values supported homoscedasticity. Additionally, random effects for participants and questions showed substantial variance (0.3789 and 0.3587, respectively, with a residual variance of 1.0584). Across all questions, Neura achieved a significantly higher normalized score of 86.17% versus 55.11% for neurologists (SD = 14.81, range = 30.85-80.85; averages of 66.38% for residents and 48.07% for board-certified physicians) — (Estimate = 1.46, Std. Error = 0.39, df = 129, t = 3.75, p < 0.001, linear mixed-effects model, and, F(1, 129) = 14.021, p < 0.001, type III ANOVA). For differential diagnosis questions, Neura achieved a normalized score of 85% versus 46.15% for neurologists (SD = 15.24, range = 26.67-78.33; averages of 58.45% for residents and 39.40% for board-certified physicians). For final diagnosis, Neura achieved a normalized score of 88.24% versus 70.93% for neurologists (SD = 17.36, range = 35.29-97.06; averages of 80.5% for residents and 64.87% for board-certified physicians) (Figure 2). The mean number of null scores and question failures was 2 and 0 for Neura and 2 and 0.4615 for neurologists (1 and 0.2 for residents and 2.625 and 0.625 for board-certified physicians). Neura obtained 15 maximum scores (p < 0.001, MCS) in its 20 evaluations, 6 of the 8 highest scores (p < 0.001, MCS), and 4 of the 11 preferred answers from both evaluators (p = 0.03, MCS) (Figure 3). In comparison, the best neurologist, a resident, obtained a normalized score of 80.85%, with 9 maximum scores, including 2 highest scores from one evaluator without a preferred answer and one null score. Neurologists’ mean response times for differential and final diagnosis were 9.62 (SD = 4.47, range = 4-32) and 8.85 minutes (SD = 5.53, range = 1-30), compared to Neura’s mean times of 0.48 and 0.317 minutes, respectively. All references provided by Neura were classified as relevant, and the information generated was deemed accurately derived from the cited sources, with no instances of hallucinatory content detected.

**Figure 2.**
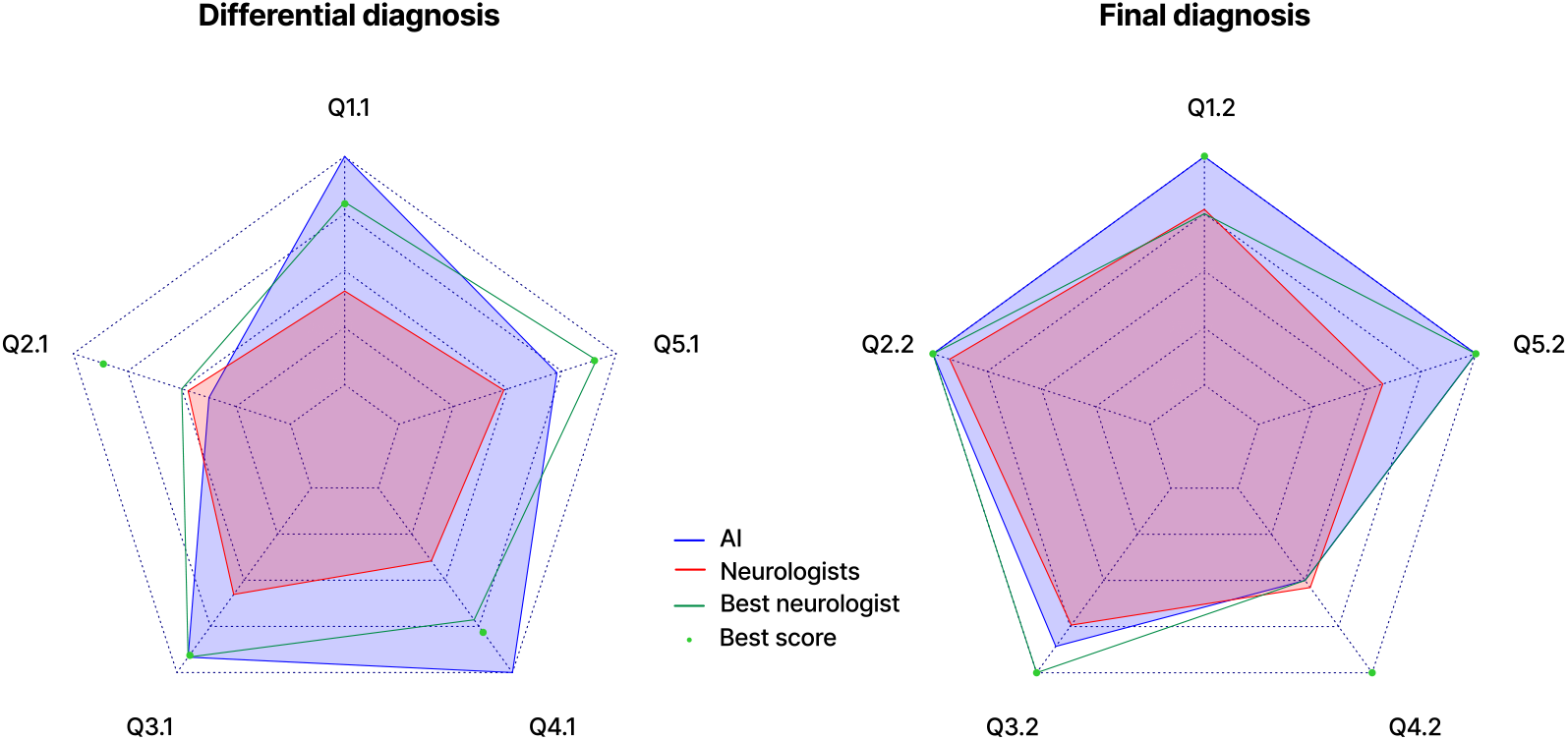
Radar charts of the performance across the differential diagnosis and final diagnosis challenges for Neura, all neurologists collectively (‘Neurologists’), the best-performing individual neurologist (‘Best neurologist’), and the best individual score from all neurologists for each question (‘Best score’). Each axis corresponds to a specific question, designated as Qx.y (where ‘x’ is the case number and ‘y’ is the tier), with scores normalized and depicted in increments of 25%.

**Figure 3.**
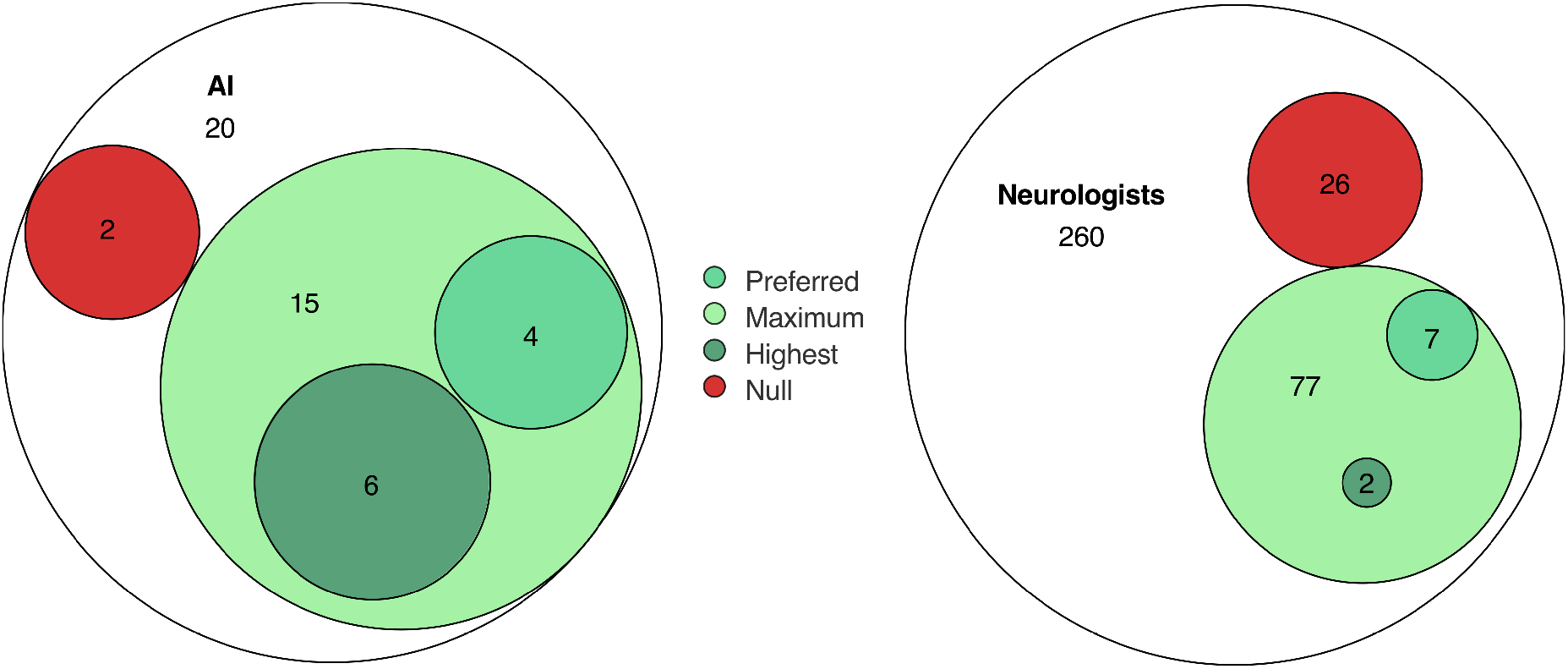
Euler diagrams representing Neura and neurologists’ answer attributes, showing total and respective counts and proportions of evaluations in each category — null scores, maximum scores, highest scores, and preferred answers.

**Figure 4.**
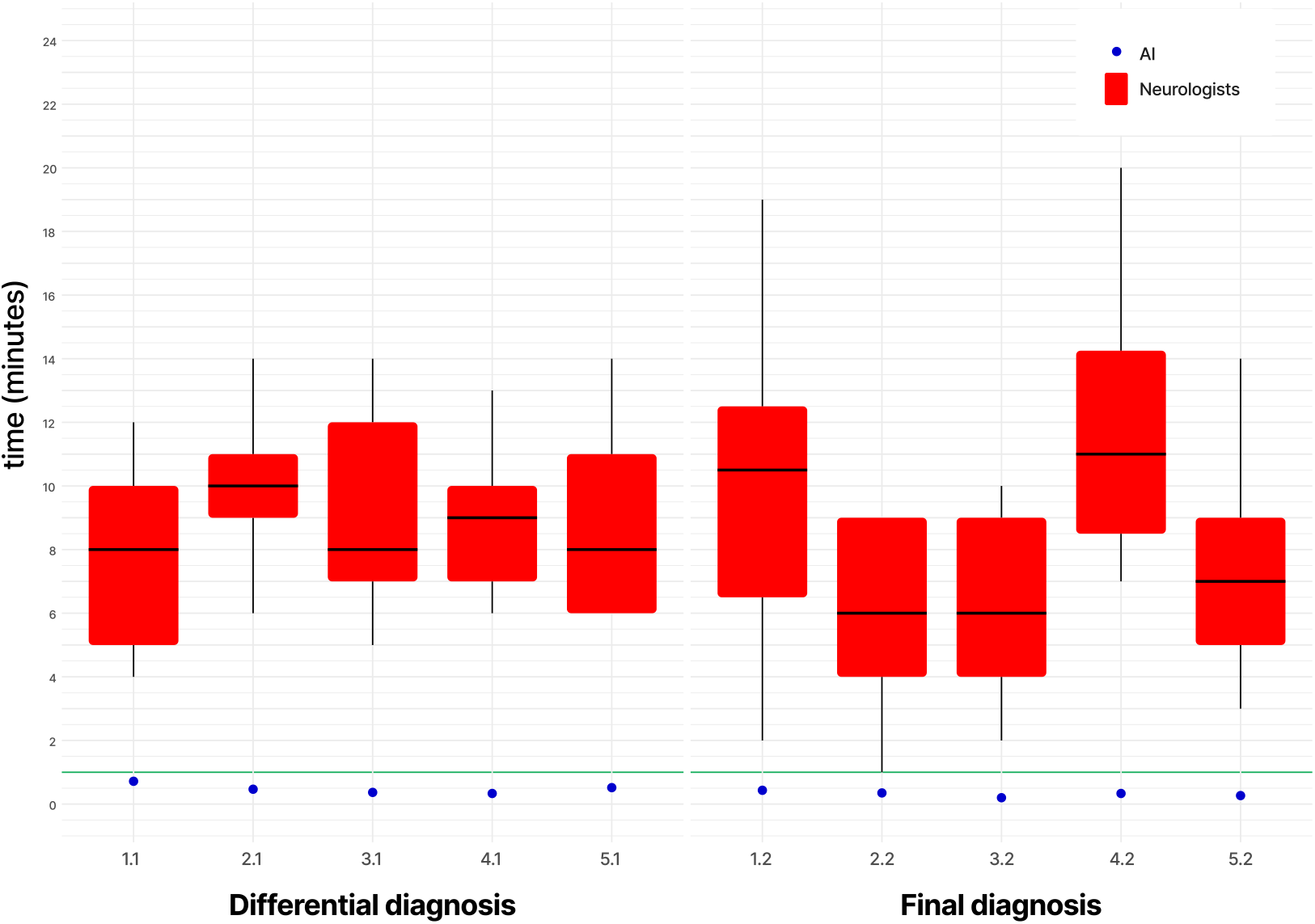
Box plots displaying the distribution of neurologists’ response times, with Neura’s times as distinct points —horizontal green lines mark a one-minute reference.

## 4 Conclusion

We introduced a scalable solution to specialize LLM in research and medicine, harnessing it to a curated corpus of knowledge. Blindly evaluated in a naturalistic setting, this solution demonstrated diagnostic acumen, perspicuity, and cogency on a select set of complex clinical cases compared to a cohort of neurologists. In line with XAI, we confirmed its controllability and reliability by ascertaining its provision of verifiable, relevant, and accurate information.

## Data Availability

All data produced in the present study are available upon reasonable request to the authors.

## Acknowledgements

We express our gratitude to *Fondation de l’Avenir* and *Académie Nationale de Chirurgie*, particularly Prof. Jean-Jacques Lemaire, Mrs. Marion Lelouvier, Dr. Ingrid Zwaenepoel, Prof. Pascal Rischmann, Dr. Hubert Johanet, Prof. Albert-Claude Benhamou, and Dr. Jean-Claude Couffinhal for their unwavering support.

